# Strong and Significant Associations of Single Nucleotide Variants (SNVs) in the Promoter and 3’-Untranslated Region (3’-UTR) of *Vascular Endothelial Growth Factor* (*VEGF*) Gene with Head and Neck Cancers

**DOI:** 10.1101/2021.03.02.21252743

**Authors:** Sadia Ajaz, Rabbia Muneer, Aisha Siddiqa, Muhammad Ali Memon, Sadaf Firasat, Aiysha Abid, Shagufta Khaliq

## Abstract

**Background:** Vascular Endothelial Growth Factor (VEGF) has a potent role in tumorigenesis and metastasis. However, data for the role of common single nucleotide variants (SNVs) in the highly polymorphic *VEGF* gene in head and neck cancers (HNCs) is limited in general and unavailable in South Asian populations.

The present study addresses this shortfall. It investigates the association of two *VEGF* SNVs, −2578 C/A (rs699947) in the promoter region and +936 C/T (rs3025039) in 3’-UTR, with the risk of HNCs and tumour characteristics.

**Methods:** The study comprised 323 participants with 121 HNC patients and 202 controls. Germline DNA was isolated from peripheral blood samples. PCR-RFLP methods were optimized and validated by Sanger sequencing. After Hardy-Weinberg evaluation, the independent associations were analyzed by applying genetic models. The χ^2^ test of independence or Fisher’s Exact test (significant p-values at <0.05) were performed and ORs (odds ratios) with 95% confidence interval were tabulated.

**Results:** *VEGF* −2578 A-allele, A-carrier and AA genotypes had significant protective association against HNCs. The respective ORs were: 0.651 (0.469 – 0.904), 0.613 (0.381 – 0.985), and 0.393 (0.193 – 0.804). *VEGF* +936 T-allele, CT and T-carrier genotypes had significantly increased susceptibility for HNCs. The respective ORs were 1.882 (1.001 – 3.536), 2.060 (1.035 – 4.102), and 2.023 (1.032 – 3.966). Additionally, *VEGF* +936 CT and T-carrier genotypes showed significant associations with higher tumour grade (p-value <0.029, and <0.037, respectively).

**Conclusion:** The present study is the foremost report of independent and unique associations of the investigated *VEGF* SNVs with HNCs.

## INTRODUCTION

VEGF is a potent mitogen and a critical pro-angiogenic factor [1–3]. Increased VEGF expression is associated with tumourigenesis [4, 5] and metastasis [6]. VEGF concentrations, and hence the function, may vary depending upon the sequence variations in the encoding gene [7].

The *VEGF* gene, is located on chromosome 6p21.1. It is 16kb in size. It is one of the most polymorphic genes with more than 100 reported genetic variations [8]. *VEGF* gene expression variations have been shown in association with the following SNVs: −1154 G/A (rs1570360), – 2578 C/A (rs699947) and −1498 C/T (rs833061) polymorphisms in the promoter region, −634 G/C (rs2010963) in the 5’ untranslated region (UTR) and +936 C/T (rs3025039) in the 3’ UTR [7, 9–10]. These SNVs have been associated with certain epithelial cancers in different populations.

Head and neck cancers (HNCs) show high incidence in South-Asia [11]. However, no data is available for the contribution of variations in critical genes like *VEGF* to HNCs in this region. In order to address the paucity of data, the present study performs the association analysis of two frequent SNV in the *VEGF* gene, rs699947 (or −2578 C/A) and rs3025039 (or +936 C/T) with HNC cases from Pakistan.

## MATERIALS AND METHODS

### Ethics Statement

The protocol is approved by the ethical review committees of the participating institutions. These include the independent ethics review committee (IEC) of the International Center for Chemical and Biological Sciences (ICCBS), University of Karachi, Karachi, Pakistan [ICCBS/IEC-016-BS/HT-2016/Protocol/1.0], the Ethics Review Committee (ERC) of the Atomic Energy Medical Centre (AEMC), Jinnah Postgraduate Medical Centre (JPMC), Karachi, Pakistan [Admin-3 (257)/2016], and the ERC, Sindh Institute of Urology and Transplantation (SIUT), Karachi, Pakistan. All the participants signed a written informed-consent form prior to sampling.

### Study Population, Demographic, and Tumour Data Collection

The present study comprised two components: case-control study design to assess the contribution of selected SNVs to HNC susceptibility in the local population, and a cross-sectional study design to evaluate the association of particular SNVs with tumour progression. The participants belonged to Southern-Pakistan. The sampling for cases was carried out at AEMC, JPMC, Karachi, Pakistan from July 2016 – July 2017, while the samples from controls had been collected earlier at SIUT, Karachi, Pakistan from March 2009 – November 2012.

All cases were biopsy-proven primary HNC patients. Information regarding tumour site, histopathology, tumour size, tumour grade, and TNM classification were recorded from the pathology reports in the patients’ medical files. For controls, blood samples from individuals in the age range of 40 – 60 years were selected. In particular, the controls visited SIUT and had no personal history of any cancer at the time of sampling as determined from their medical records.

### Sample Collection

The present study comprised 323 participants, with 121 cases and 202 controls. In case of patients 8-10ml, while for controls 4-5ml of blood samples were collected in ACD coated vacutainers and either processed immediately or stored at 4*°*C until DNA extraction.

### DNA Extraction and Spectrophotometric Analysis

DNA was extracted using standard phenol-chloroform method [12]. Quality of the extracted DNA and concentration were determined spectrophotometrically. The samples had OD_260_/OD_280_ ratio between 1.7 – 1.9. No DNA fragmentation was observed in agarose gel run for quality control purpose

### Genotyping

Genotyping for *VEGF* −2578C/A (rs699947) and +936C/T (rs3025039) were carried out by <u>P</u>olymerase <u>C</u>hain <u>R</u>eaction (PCR) followed by <u>R</u>estriction <u>F</u>ragment <u>L</u>ength <u>P</u>olymorphism (RFLP) at Dr. Panjwani Center for Molecular Medicine and Drug Research (PCMD), ICCBS, University of Karachi, Karachi, Pakistan.

Genotyping accuracy was validated by sequencing selected samples for the detected polymorphic variants by Sanger sequencing, commercially.

PCR-RFLP based genotyping assays have been described previously [13, 14].

Genotyping was confirmed by second blind evaluation. Overall evaluation of concordance rates was 100% for both *VEGF* −2578C/A and +936C/T polymorphisms. The call rates of genotyping reproducibility were also 100% for randomly selected samples.

### Validation of Genotyping Assays

The genotyping assays were further validated by Sanger sequencing commercially.

## Statistical Analysis

The demographic, clinical, and laboratory data were entered, coded, and processed with SPSS^®^ software (version 19). The selected SNPs were tested for Hardy-Weinberg equilibrium (HWE) [14]. The p-values >0.05 showed that the genotypes were in HWE.

Associations between each SNP, HNC risk, and clinico-pathological features were analyzed by Chi-squared test of independence or Fisher’s exact test, where appropriate, with requisite degrees of freedom. Statistical significance was selected at p-values < 0.05. Odds ratios with 95% confidence interval were calculated to quantify the strength of genetic association.

## RESULTS

### Demographic Comparison and Clinical Data

The demographics of 121 cases and 202 controls were analyzed. The mean ages of cases and controls were 50.06 ± 1.29 and 46.36 ± 0.36, respectively

The clinico-pathological features of the patients are presented in table 1.

**Table 1.** Histopathological features of the HNC tumours of the cases.

| **S.No** | **Parameter\*** | **N or value %** |
| --- | --- | --- |
| **1** | **Gender (n=108)** |  |
| **1a** | Male | 75 (69.4%) |
| **1b** | Female | 33(30.6%) |
| **2** | **Tumor Site (n=100)** |  |
| **2a** | Buccal cavity/oral cavity | 38(38%) |
| **2b** | Cheek | 19(19%) |
| **2c** | Tongue | 14(14%) |
| **2d** | Larynx | 8(8%) |
| **2e** | Pharynx | 7(7%) |
| **2f** | Lip | 5(5%) |
| **2g** | Unspecified parts of mouth | 5(5%) |
| **2h** | Nasal cavity | 2(2%) |
| **2i** | Glottis | 1(1%) |
| **2j** | Salivary gland | 1(1%) |
| **3** | **Histological Sub-type (n=102)** |  |
| **3a** | SCC | 92(90.2%) |
| **3ai** | MD SCC | 45(44.11%) |
| **3aii** | WD SCC | 21(20.6%) |
| **3aiii** | PD SCC | 7(6.86%) |
| **3aiv** | Well to MD SCC | 6(5.88%) |
| **3b** | Sarcomas | 3(2.95%) |
| **3c** | Basal cell carcinoma | 2(1.96%) |
| **3d** | Adenoid cystic carcinoma | 2(1.96%) |
| **3e** | Others | 2(1.96%) |
| **3f** | Acinic cell carcinoma | 1(0.98%) |
| **4** | **Tumor size (n=80)** |  |
| **4b** | □5cm | 48(60%) |
| **4a** | ≥5cm | 32(40%) |
| **5** | **Tumor stage (n=74)** |  |
| **5a** | Stage (III-IV) | 61(82.44%) |
| **5b** | Stage (I-II) | 13(17.56%) |
| **6** | **Tumor Grade (n=61)** |  |
| **6a** | Low grade (1-2) | 56(91.8%) |
| **6b** | High grade(3-4) | 5(8.2%) |

SCC=Squamous cell carcinoma, MD SCC= Moderately differentiated Squamous cell carcinoma, WD SCC= Well differentiated Squamous cell carcinoma, PD SCC= Poorly differentiated Squamous cell carcinoma
\*missing data

### Significant Protective Effect of *VEGF* (rs699947) −2578 A-Allele, A-carrier Genotype, and AA Genotype Against HNCs in Dominant, Recessive and Co-Dominant Multiplicative Models

The representative gels for amplification and restriction digestion are shown in Figures 1 and 2, respectively. Validation by Sanger sequencing is shown in the inset of figure 2.

**Figure 1.**
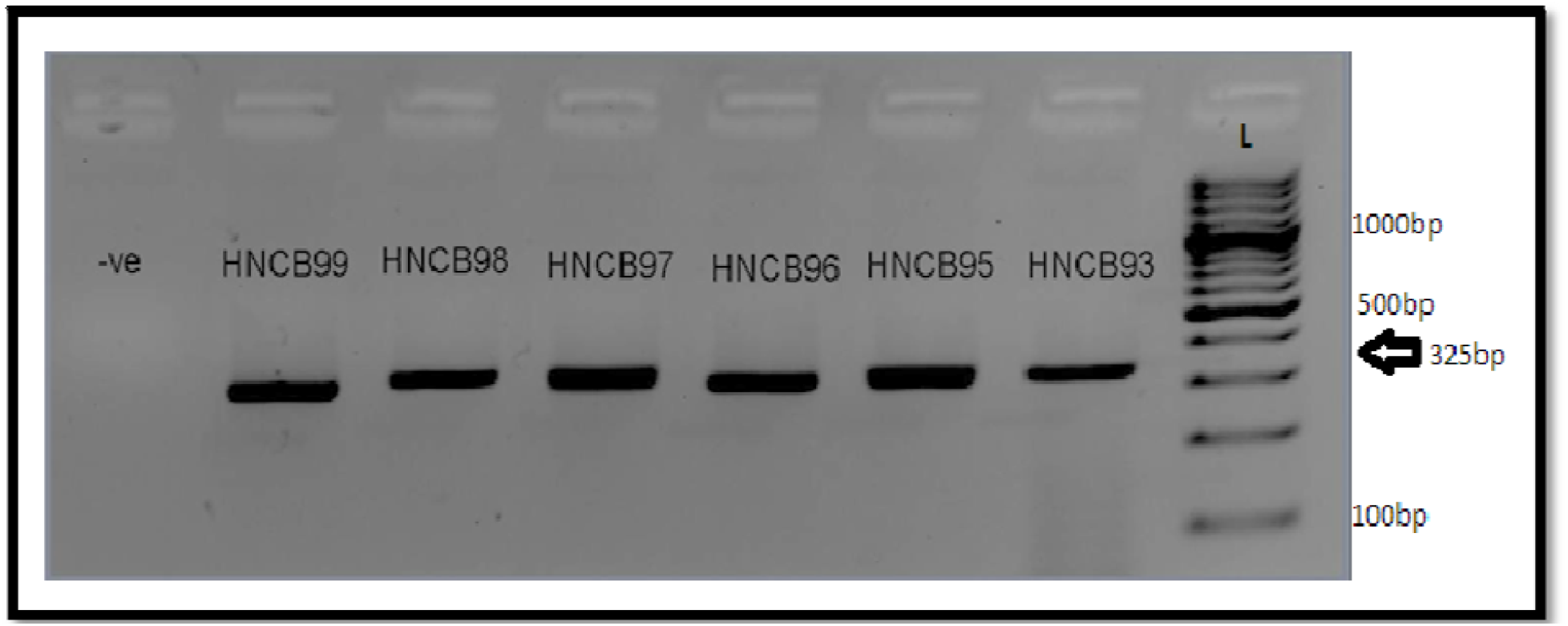
Amplification of *VEGF* −2578C/A on 2.5% agarose gel with 100 bp ladder as a DNA marker. Ethidium bromide was used for staining.

**Figure 2.**
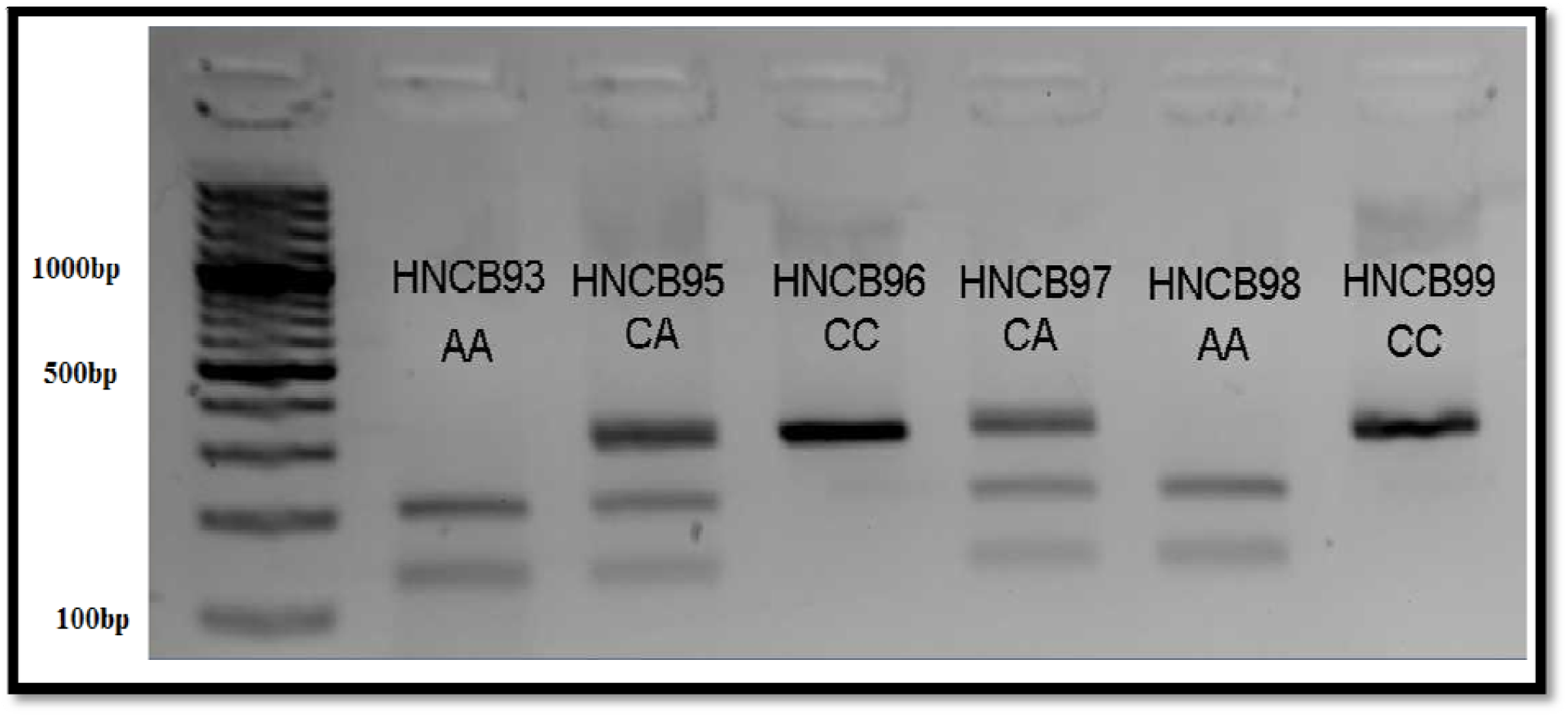
Genotyping of *VEGF* −2578C/A digested with *BglII* enzyme. 325 bp DNA band showed CC genotype, combination of 123 bp, 202 bp, and 325 bp DNA band showed CA genotype, while 202 bp and 123 bp DNA band represented AA genotype.100 bp ladder was used as a marker.

**Figure 2.**
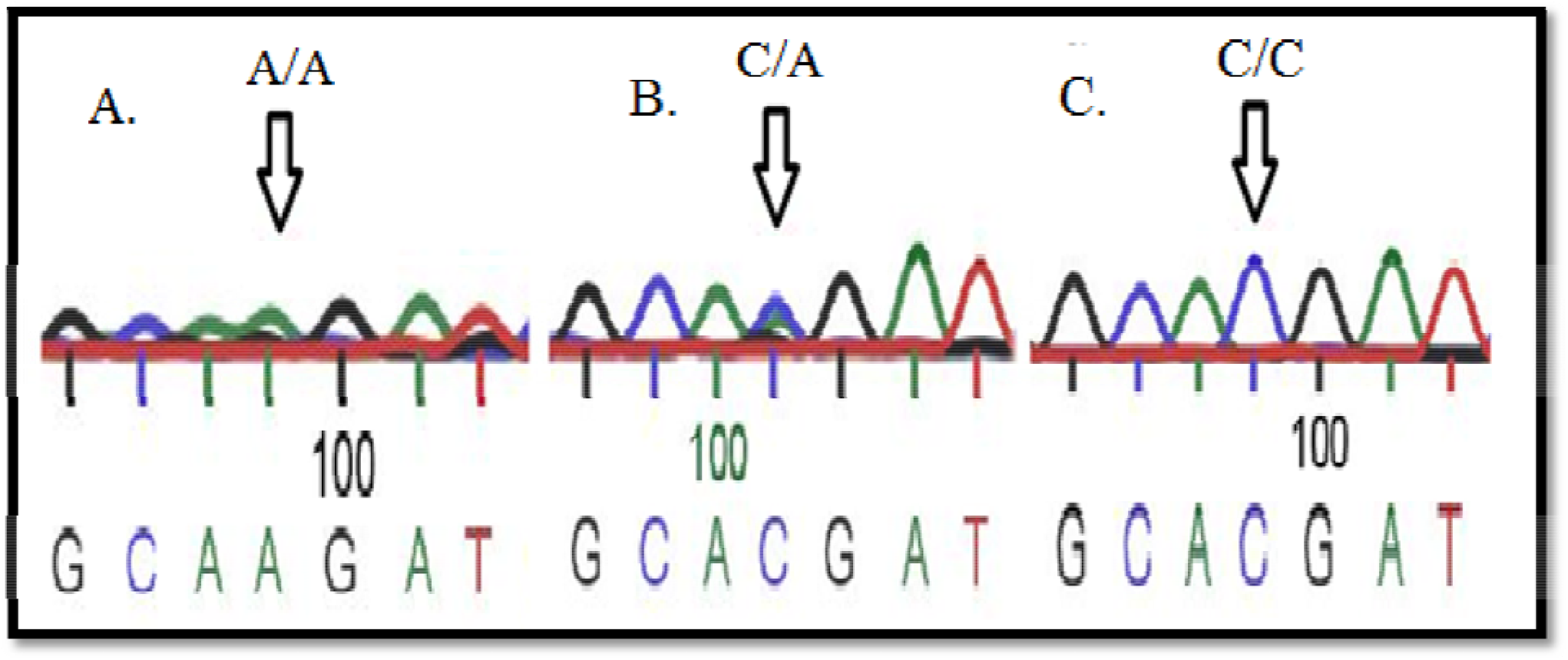
(inset): Electropherogram of Sanger sequencing for *VEGF* −2578C/A polymorphism (arrows represent the site of polymorphism). A-Variant homozygous (AA), B-Heterozygous variant (CA), C-Wildtype homozygous (CC).

The genotypes for *VEGF* −2578C/A polymorphism (CC, CA, and AA) were in Hardy-Weinberg equilibrium for both cases and controls (HNC patients: χ^2^=0.13, 2 df, *p* >0.05; Controls: χ^2^=0.6, 2 df, *p* >0.05).

The genotypic and allelic frequencies of *VEGF −*2578C/A polymorphism are given in the table 2. As shown in the table, a significant protective effect of A-allele (OR.: 0.65; 95%CI: 0.47 – 0.90), A-carrier and AA genotypes [OR.: 0.61 (95% CI: 0.38 – 0.98; OR: 0.39 (95% CI 0.19 – 0.80)] were observed against HNCs.

**Table 2.** Distribution of the *VEGF* −2578C/A genotypes and allele frequencies (with standard errors) in controls and cases (HNC patients). Chi-squared test and odds-ratios (95% Confidence Interval) for the risk of HNC.

| *VEGF* -2578C/A Polymorphism | Controls (n = 202) | HNC patients (n = 121) | Chi-square (p-value) | Odds-ratios (OR) (95% Confidence Interval) |
| --- | --- | --- | --- | --- |
| **Genotypes** | N (%) | N (%) |  |  |
| CC | 58 (28.7%) | 48 (39.7%) | Ref. | Ref. |
| CA (Over-Dominant Model) | 101 (50% %) | 59 (48.8%) | 1.88 (0.17) | 0.71 (0.43– 1.16) |
| A-Carrier (Dominant Model) | 144 (71.3%) | 73 (60.3%) | 4.12 (0.04)* | 0.613 (0.381 – 0.985)* |
| AA (Recessive Model) | 43 (21.3%) | 14 (11.6%) | 6.75 (0.009)** | 0.393 (0.193 – 0.804)** |
|  |  |  | Co-dominant Multiplicative Model |  |
| **Allele Frequencies** |  |  |  |  |
| p[C] | 0.54±0.04 | 0.64±0.04 | Ref. | Ref. |
| q[A] | 0.46±0.04 | 0.36±0.04 | 6.62 (0.01)** | 0.651 (0.469 - 0.904)* |

\*significant; \*\*highly significant

### Significant Susceptibility Risk of *VEGF* (rs 3025039) +936 T-Allele, CT, and T-carrier Genotypes with HNCs in Dominant, Over-Dominant and Co-Dominant Additive Model

The representative gels for amplification and restriction digestion are shown in Figures 3 and 4, respectively. Validation by Sanger sequencing is shown in the inset of figure 4.

**Figure 3.**
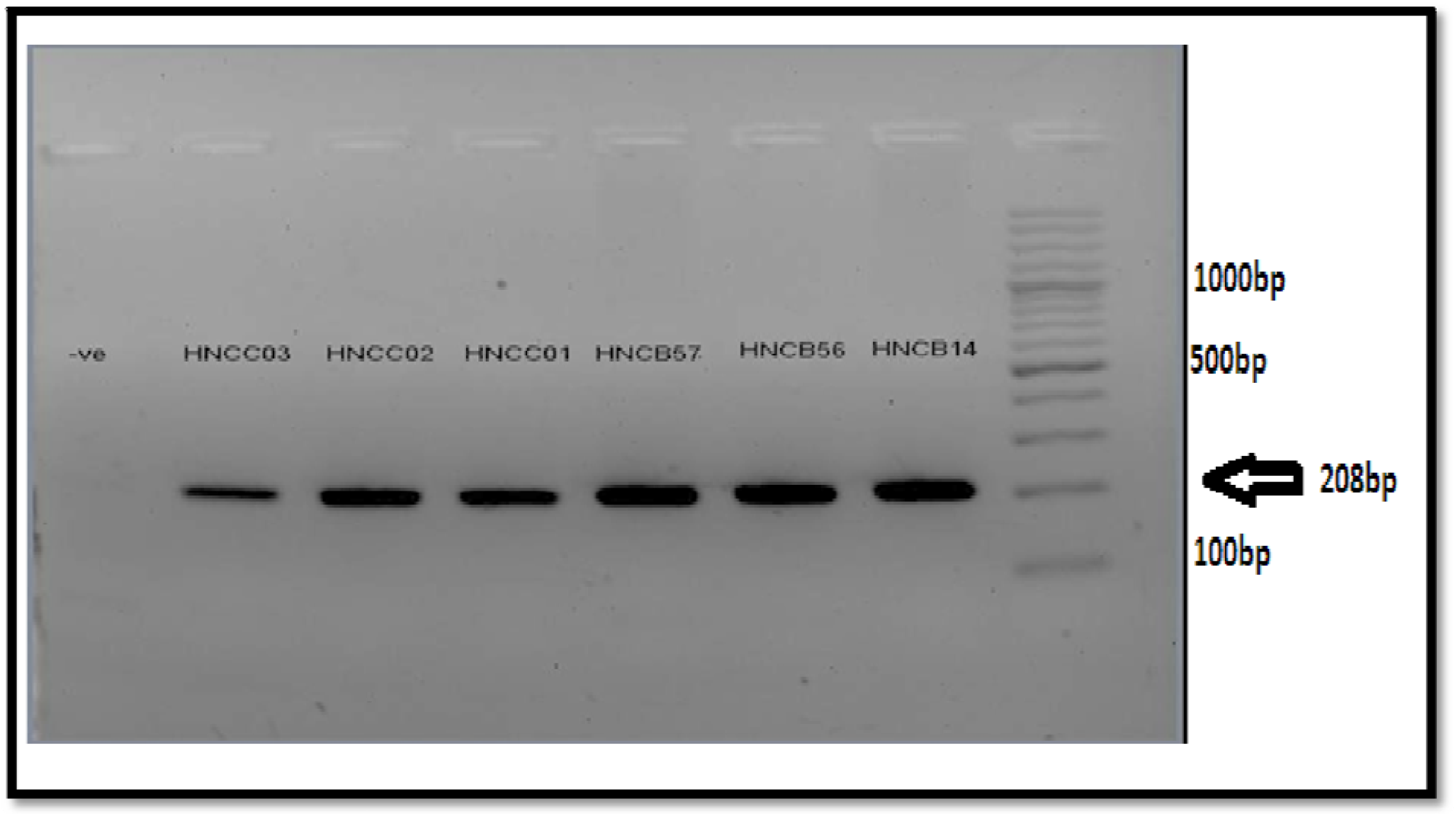
Amplification of *VEGF* +936C/T on 2.5% agarose gel with 100 bp ladder as a DNA marker. Ethidium bromide was used for staining.

**Figure 4.**
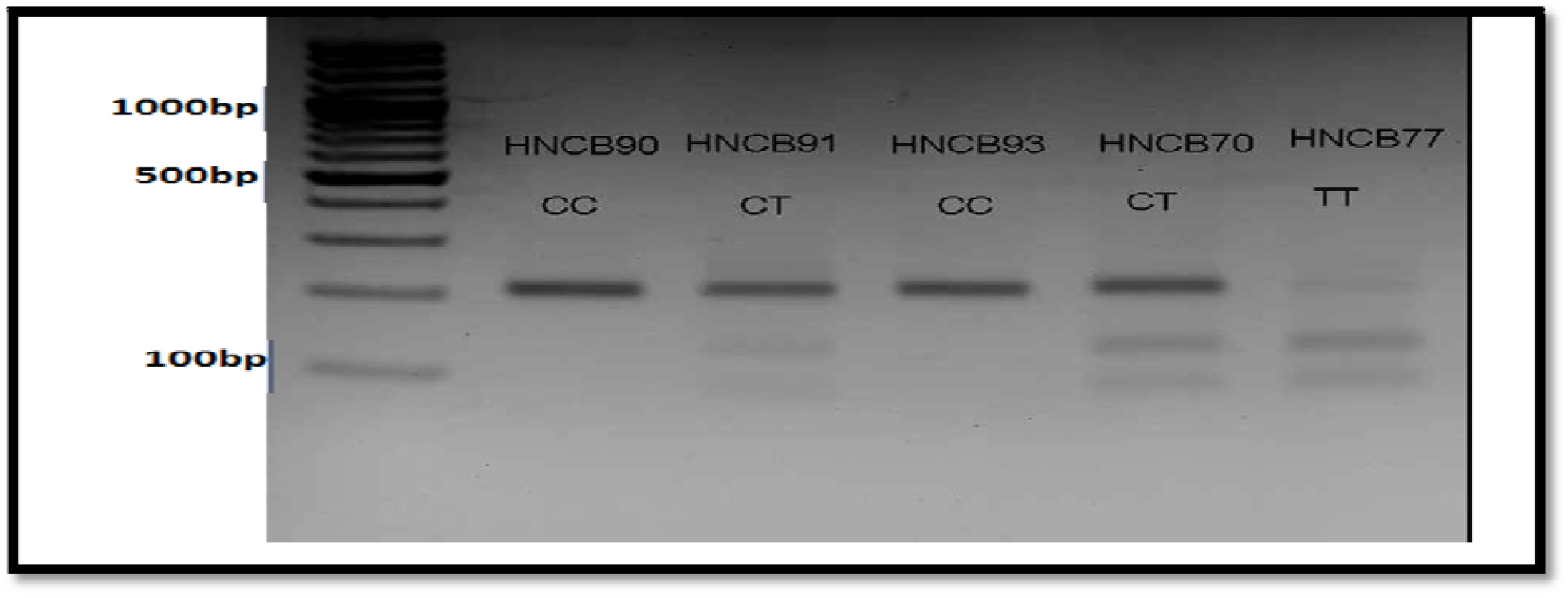
Genotyping of *VEGF* +936C/T digested with *NlaIII* enzyme. A-DNA band of 208 bp represented CC genotype, while 86 bp, 122 bp, and 208 bp DNA bands showed CT genotype, B-122 bp and 86 bp DNA band depicted TT genotype.100 bp ladder was used as a marker.

**Figure 4.**
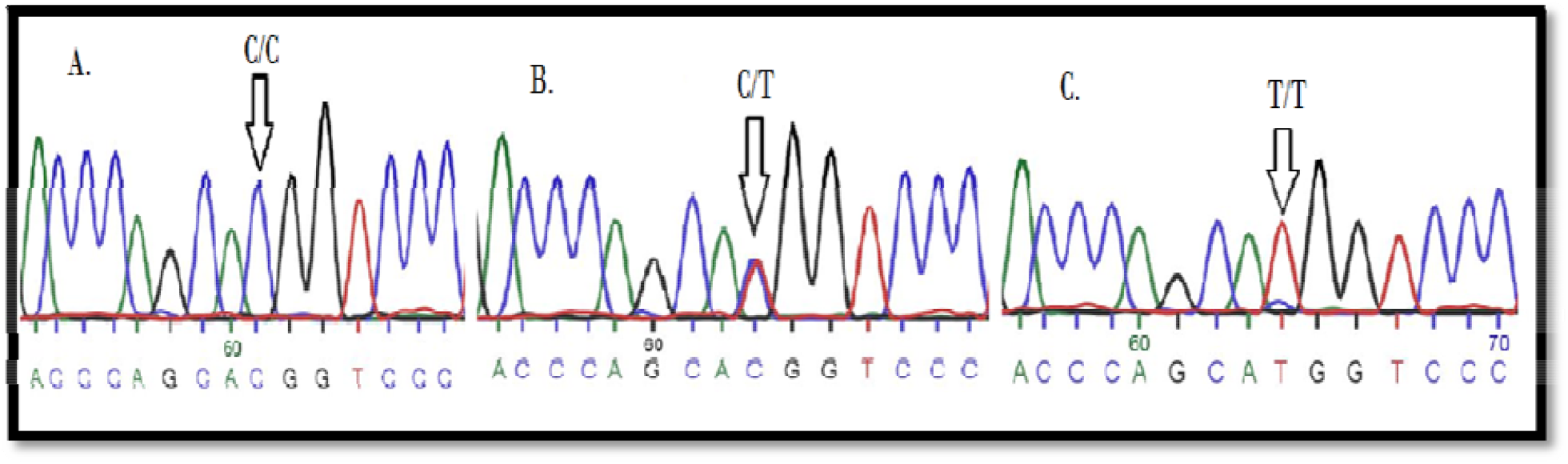
(inset): Electropherogram of Sanger sequencing for *VEGF* +936C/T SNP (arrows represent the site of polymorphism). A-Wildtype homozygous (CC), B-Variant heterozygous (CT), C-Variant homozygous (TT).

The genotypes for *VEGF* +936C/T polymorphism (CC, CT, and TT) were in Hardy-Weinberg equilibrium for both cases and controls (HNC patients: χ^2^=0.08, 2 df, *p* >0.05; Controls: χ^2^=0.5, 2 df, *p* >0.05).

The genotypic and allelic frequencies of *VEGF* +936C/T polymorphism are given in the table 3. The chi-squared test of independence showed that *VEGF* +936C/T polymorphism was associated with the risk for HNCs. A significantly increased risk for HNCs was observed for T-allele, CT and T-carrier genotypes in the analyzed case-control group.

**Table 3.** Distribution of the *VEGF* +936C/T genotypes and allele frequencies (with standard errors) in controls and cases (HNC patients). Chi-squared test and odds-ratios (95% Confidence Interval) for the risk of HNC.

| *VEGF* +936C/T Polymorphism | Controls (n = 156) | HNC patients (n = 121) | Chi-square (p-value)/Fisher's Exact Test (p-value) | Odds-ratios (OR) (95% Confidence Interval) |
| --- | --- | --- | --- | --- |
| **Genotypes** | N (%) | N (%) |  |  |
| CC | 139 (89.1%) | 97 (80.2%) | Ref. | Ref. |
| CT (Over-dominant Model) | 16 (10.3%) | 23 (19.0%) | 4.35 (0.037)* | 2.06 (1.03– 4.10)* |
| T-Carrier (Dominant Model) | 24 (10.9%) | 24 (19.8%) | 4.32 (0.038)* | 2.02 (1.03 – 3.96)* |
| TT (Recessive Model) | 01 (0.6%) | 01 (0.8%) | 0.06 (1.00)<sup>†</sup> | 1.43 (0.09 – 23.19) |
|  |  |  | Co-dominant Additive Model |  |
| **Allele Frequencies** |  |  |  |  |
| p[C] | 0.94±0.02 | 0.87±0.03 | Ref. | Ref. |
| q[T] | 0.06±0.02 | 0.13±0.03 | 3.96 (0.047)* | 1.882 (1.001– 3.536)* |

\*significant result; <sup>†</sup>Fisher's Exact Test;

### Significant Association of *VEGF* +936 CT, and T-carrier Genotypes with Tumour Grade in HNCs

The association analysis of *VEGF* −2578C/A and +936C/T polymorphism with clinico-pathological features of HNC cases is shown in table 4. Significant associations were observed between CT- and T-carrier genotypes with tumour grade in HNCs. The odds ratios (95% CI) were 10.29 (1.45 – 72.81) and 9 (1.29 – 62.59), respectively.

**Table 4.** Chi-squared test for the association of −2578 C/A and +936 C/T alleles with tumour characteristics in HNCs.

| **VEGF**<br>**POLYMORPHISMS** | **Tumour Size**<br>**(≤5 vs >5)**<br>**p-value** | **Tumour Stage (T1-**<br>**T2 vs T3-T4)**<br>**p-value** | **Tumour Grade (G1-G2 vs G3-G4)**<br>**p-value** |
| --- | --- | --- | --- |
| -2578C (AA vs AC/CC) | 0.722 | 0.892 | 0.792 |
| -2578C (AA vs CC) | 0.615 | 0.039 | 0.61 |
| -2578C (AC vs CC) | 0.42 | 0.471 | 0.459 |
| **+936 (CC/CT vs TT)** | 0.604 | 0.358 | **0.037\*\*** |
| +936 (CC vs TT) | 0.09 | 0.6 | 0.838 |
| **+936 (CC vs CT)** | 0.652 | 0.246 | **0.029\*\*** |

\*\*statistically significant

## DISCUSSION

Cancer is a complex disorder. It has a number of etiologies. The underlying causes could be environmental (e.g. UV exposure), occupational (e.g. chemicals, radioactive materials and asbestos), life style related factors (e.g. tobacco, alcohol) and biological factors including age or inherited genetic defects. In the last group, VEGF plays an important role in tumour development and progression among different epithelial cancers [16].

Genetic variations in the regulatory region and/or the gene itself influence gene expression. Thus, these variations are potential genetic markers in disease susceptibility, progression, and treatment response [17]. In the highly polymorphic *VEGF* gene, SNVs in the regulatory regions, i.e. promoter, 5’ UTR, and 3’ UTR have been linked with variations in the gene expression [16, 18–19]. These polymorphisms have been associated with carcinomas including breast [20], melanoma [21], lung [22], kidney [13], and prostate [23] cancers.

Here, we report significant associations of two SNVs in the *VEGF* gene, −2578C/A in the promoter and +936C/T in 3’ (UTR), with HNCs. To the best of our knowledge, this is the first such report from South Asia.

Among the eligible published reports from other parts of the world (i.e. studies which fulfill STROBE criteria) [24], two studies each have reported association of *VEGF* −2578 C/A [25, 26] and +936 C/T with HNCs risk [25, 27].

The present study reports strong and significant protective effect of −2578 A-allele, A-carrier, and AA genotypes against HNCs. The earlier published reports are from Caucasian populations, one from Germany [25] and the other from Serbia [26]. In contrast to the present study, the German study reports increased risk for oral squamous cell carcinomas (OSCC) with −2578CA genotype, while Serbian study does not report independent association of this SNV with HNCs. It is pertinent to mention here that the power of the present study for *VEGF* −2578C/A is more than 90%.

In 3’ UTR, +936C/T was not linked with risk for OSCCs in Taiwanese population [27]. However, in the German study +936 T allele was associated with increased risk [25], which is in agreement with the present report. The frequency of T allele in Caucasian populations is much higher (approx. 23%) than the South-Asian populations (approx. 10%) [9]. This frequency is further reduced to 7% in PJL (Punjabis in Lahore)-1000 genomes project, and the present study. Mechanistically, the role of VEGF polymorphisms in modulating the VEGF expression has been reported with varying results [28, 29]. For *VEGF* −2578C/A, the correlation of AA genotype with lower VEGF levels has been shown experimentally [30,31]. In agreement, another study reports that C-allele is associated with higher VEGF levels [32]. In contrast one study reports the co-relation of A allele to higher VEGF mRNA levels [33].

The possible mechanism for the observed association may be explained by the binding of two transcription factors, GATA-2 and MZF-1 to the promoter region of *VEGF* gene.

The −2578C/A SNP is positioned in the GATA-2 binding site. The A allele is likely to reduce the binding specificity. It is proposed that the −2578 C allele produces a binding site for GATA-2, that ultimately permits activation of *VEGF* gene. Therefore, the −2578 C allele may increase VEGF levels and promotes angiogenesis as well as tumor cells proliferation. Whereas, the −2578 A allele can reduce the transcription by decreasing the binding affinity to this region [34]. *VEGF* promoter region also has a binding site for the Myeloid Zinc Finger-1 (MZF-1), which is likely to regulate *VEGF* expression [35].

Additionally, it is reported that −2578 C/A polymorphism in *VEGF* promoter region has association with −2549 position, which has an18 bp fragment deletion/insertion (D/I) polymorphism. In the previous study it was shown that individuals with the −2578A allele had 18 bp insertion, while those with the −2578C allele did not have this insertion (−2549 D). It is also suggested that the linkage disequilibrium between these two sites is correlated with increased transcriptional activity [35].

Among different cancers, various studies have reported the contribution of *VEGF* gene expression with conflicting results in oral cancers. Still, a substantial number of studies have reported no association between *VEGF* expression and angiogenesis in oral cancers [34]. In Tunisian population, *VEGF* −2578C/A has been correlated with nasopharyngeal carcinomas (NPCs), where C allele was associated with NPCs in this population [34]. In Caucasian population, this polymorphism has correlation with breast cancer [35]. While, in Pakistani population the −2578 A allele has significant association with increased risk of renal cell carcinomas [13], in contrast to the present study, which reports a protective effect in HNCs.

In the present study, T allele in *VEGF* +936C/T polymorphism has been associated with higher risk for HNCs. However, T allele has been associated with lower VEGF levels [38–39]. An AP-4 (<u>A</u>ctivator <u>E</u>nhancer-<u>B</u>inding <u>P</u>rotein 4) DNA binding site (CAGCTG) is present in the 3′-UTR of *VEGF*. The C allele at +936 has the binding site for this protein, which is absent in case of +936 T allele [40], ultimately affect *VEGF* expression [41].

To address these discrepancies, we propose, that the allele could be in linkage disequilibrium with another locus, which increases the risk for HNCs. Indeed, +1451 C/T (rs3025040) in 3’UTR, located in the miR-199a or miR-199b binding site exhibits strong linkage disequlibrium (r^2^ = 1.0) with +936C/T in Chinese population [37].

Most of the published literature presents the association of these polymorphisms with tumour properties such as invasion, stage or grade. The present study also reports significant association of the polymorphism in 3’UTR with tumour grade.

Furthermore, the inconsistencies between the polymorphisms and serum concentration levels may arise from differences in the health and disease status of the participants. Thus, we recommend that well designed studies, clearly describing the characteristics of participants should be carried out.

In conclusion, to the best of our knowledge, the present study is the first report of strong and significant association of *VEGF* −2578 C/A and +936 C/T polymorphisms with HNCs in South-Asian population. This molecular investigation shall aid in establishing genotypic markers for risk and prognosis of HNCs

## Data Availability

Data available on request.

## ACKNOWLEDGEMENTS

The authors would like to thank Institution core facilities (PCMD/ICCBS), AEMC, JPMC staff and all the participants of the study for their co-operation.

